# Optimizing SARS-CoV-2 molecular diagnostic using N gene target: insights about reinfection

**DOI:** 10.1101/2020.12.06.20244905

**Authors:** Raphael Contelli Klein, Mary Hellen Fabres Klein, Larissa Gomes Barbosa, Lívia Vasconcelos Gonzaga Knnup, Larissa Paola Rodrigues Venâncio, Jonilson Berlink Lima, Théo Araújo-Santos

## Abstract

**Introduction:** Molecular diagnosis of SARS-CoV-2 is a huge challenge to many countries around the world. The cost of tests to check infected people is inaccessible since specialized teams and equipment are not disposable in remote locations. Herein, we compared the fitness of two primers sets to the SARS-CoV-2 N gene in the molecular diagnosis of COVID-19.

**Materials and Methods:** The 1029 patient samples were tested to presense/abscence molecular test using in house US CDC protocol. We compared the fitness of two primers sets to two different regions of N gene targets.

**Results:** Both targets, N1 and N2 displayed similar fitness during testing with no differences between Ct or measurable viral genome copies. In addition, we verified security ranges Cts related to positive diagnostic with Ct above 35 value failuring in 66,6% after retesting of samples.

**Main conclusion:** Our data suggest that it is secure to use just one primer set to the N gene to identify SARS-CoV-2 in samples and the labs should be careful to set positive samples in high Ct values using high cutoffs.

## 1. Introduction

Demands of molecular diagnosis to testing COVID-19 are fast growing around the world and costs and efficiency of the RT-PCR technique has been in check. In this context, different RT-PCR kits are used to identify SARS-CoV-2 virus in patients’ samples are now available, using just one or multiple gene targets ^’^(Pizzol et al. 2020, Mathuria et al. 2020).

After six months in the covid-19 pandemic, Brazil is between major countries affected by disease (Hallal et al. 2020). Genome vigilance of the virus is now occurring and variants of the virus genome are disposable to genomic banks permitting check of the virus specificity of the disposable tests (Candido et al. 2020). Some studies are comparing sensitivity and specificity of different sets of probes/primers against different gene targets of the SARS-CoV-2. In this context, the conserved nucleocapsid protein gene (N) is the major target present in the disposable tests (Pizzol et al. 2020) and US CDC test includes two sets of primers with good sensitivity to virus detection (Etievant et al. 2020). In addition, recent work demonstrates some mutation in the forward primer to N gene target in Chinese CDC test, suggesting US CDC test can be more appropriate to Brazilian testing (Candido et al. 2020).

In this article, we compared the fitness between both commercial N gene targets present in the US CDC panel to SARS-CoV-2 and we identified similar results using both targets. In addition, we verified that high Ct value in patient samples presents low reproducibility, suggesting that this range of Ct value can contribute to misinterpretation in the test results.

## 2. Materials and Methods

### Clinical Samples

Nasopharyngeal, nasal and oropharyngeal swabs and sputum samples (n=1029) collected during May and June of 2020 were obtained after SARS-CoV-2 detection in the Laboratory of Vectors and Infection Disease. Residual samples were de-identified samples and considered non-human subjects of the research. These samples were used to test the fitness profile of the US CDC 2019-nCoV_N1 and 2019-nCoV_N2 primer-probe sets as described in the following sections.

### RNA isolation

Sputum and Swabs obtained from patients reporting covid-19-like synmptoms were processed to RT-qPCR SARS-CoV-2 detection. In brief, RNA isolation of samples was extracted using commercial kits following supplier’s instructions, such as PureLink® Viral RNA/DNA Mini Kit (ThermoScientific), Cellco (Cellco Biotec) and Biogene (Quibasa) and RNAs were resuspended in 60 µL of RNase-free water (GIBCO).

### RT-qPCR

Quantitative Real-time PCR analysis was performed on QuantStudio 5 Real-Time PCR system (ThermoScientific, USA) using the primer set from 2019-nCOV RUO kit (IDT Coralville, IA). The PCR reaction mixture consisted of TaqMan Fast Virus 1-Step Master Mix (ThermoScientific), 0.75 μL of primers and 2.5 μL of RNA in a final volume of 10 μL reaction. Cycling conditions were 50°C for 5 min and 95°C for 20 seconds, followed by 45 cycles at 95°C for 15 seconds and 58°C for 1 min. Alternatively, we used KAPA PROBE FAST qPCR Master Mix (2X) Kit (Sigma-Aldrich), 0.75 μL of primers and 2.5 μL of RNA in a final volume of 10 μL reaction. Cycling conditions were 42°C for 5 min and 95°C for 3 minutes, followed by 45 cycles at 95°C for 5 seconds and 60°C for 1 minute. RNAse P was used as a sample control. The primers and concentrations used in the experiment were as follows: 500 nM N1: Forward: 5′-GACCCCAAAATCAGCGAAAT-3′; 500 nM N1: Reverse: 5′- TCTGGTTACTGCCAGTTGAATCTG-3′; 125 nM N1-Probe FAM-ACCCCGCATTACGTTTGGTGGACC-NFQ-MGB; 500 nM N2: Forward: 5′- TTACAAACATTGGCCGCAAA-3′; 500 nM N2: Reverse: 5′- GCGCGACATTCCGAAGAA- 3′ and 125 nM N2-Probe FAM-ACAATTTGCCCCCAGCGCTTCAG-NFQ-MGB (“Centers for Disease Control and Prevention. 2019-novel coronavirus (2019-nCoV) real-time RT-PCR primer and probe information” 2020). Specificity of the PCR products of N1 and N2 amplification were confirmed by polyacrylamide gel electrophoresis with silver stain.

### Statistical analysis

Descriptive statistics were performed to determine the relative frequencies for categorical variables, as well as to obtain medians and their respective standard error values for continuous variables. Linear regression was built to compare N1 and N2 linearity profiles between both two targets to Ct value in all samples used in this study. A Receiver Operator Characteristics (ROC) curve analysis was performed to assess the sensibility and specificity of Ct value in a subset of samples. All data was analyzed using GraphPad Prism 5.0 (GraphPad Software Inc).

### Ethic statement

The Research Ethics Committee of UFOB approved this study in 2020 (license number: 30629520.6.0000.0008). All clinical investigations were conducted according to the Declaration of Helsinki.

## 3. Results

### No differences between N1 and N2 primers set fitness in the SARS-CoV-2 detection

Several tests use just one set of primers to detect SARS-CoV-2 (Mathuria et al. 2020). The efficiency or increment of different targets in the detection of SARS-CoV-2 was poorly addressed. Herein, we evaluate fitness of N gene targets present in UCDC test diagnosis in 1029 population naso and oropharyngeal swabs or sputum samples. We verified N1 and N2 primers sets displayed similar Ct values for each sample (Figure 1). In order to verify the fitness in each Ct value range, we quantify the frequencies of results between N1 and N2 primers sets. We verify no differences of Ct value failure to detect viral RNA between both N1 and N2 primers sets (table 1). This data suggests just one primer set could be used to test patients with SARS-CoV-2 infection potentially reducing costs during molecular testing without diminish efficiency in the diagnosis accuracy.

**Table 1.** N1 versus N2 fitness in SARS-CoV-2 detection in population.

| **Ct value group** | **Target** |  | **Specimens** |  |
| --- | --- | --- | --- | --- |
|  | **N1 %** | **N2 %** | **Sputum %** | **Swabs %** |
| **Ct undetermined** | **48,0** | **48,0** | **47,5** | **48,1** |
| **Ct <30** | **25,3** | **25,3** | **31,8** | **23,9** |
| **30 < N1 e N2 <34** | **5,4** | **5,4** | **4,5** | **5,6** |
| **Ct >34** | **10,9** | **10,9** | **9,5** | **11,2** |
| **Ct failure\*\*** | **5,1** | **5,3** | **6,7** | **11,2** |

\*Data from 1029 different patient samples.
\*\*Ct failure were just one target presents amplification

**Figure 1.**
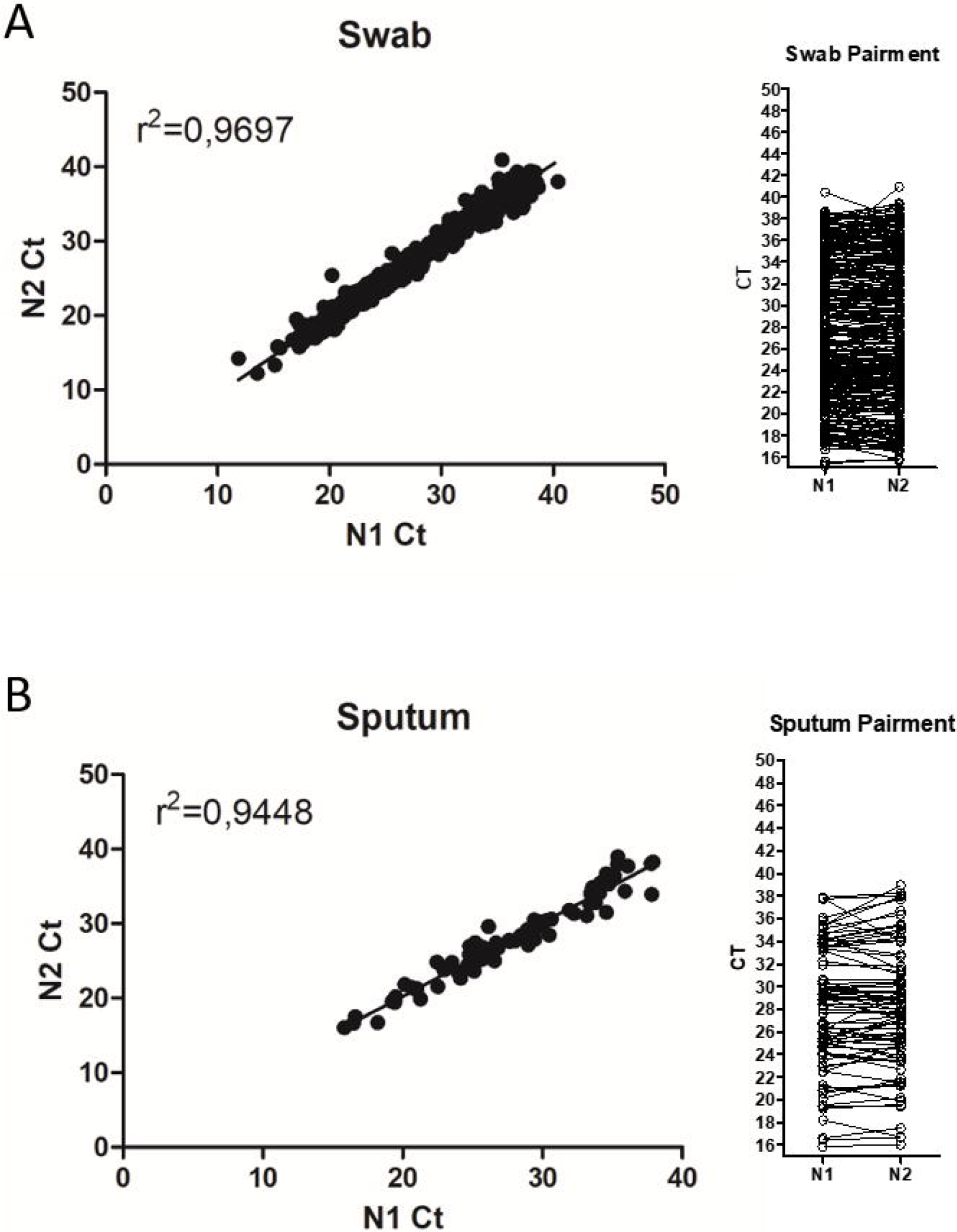
Comparison between N1 and N2 Ct values during SARS-CoV-2 detection in patient samples. Data on the graphs indicate N1 and N2 Ct values comparison for each patient samples of (A) swab and (B) sputum by linear regression (left panels) or paired analysis (right panels). p value < 0.0001 for linear regression and paired t test p < 0.9483.

### Accuracy of N gene target in SARS-CoV-2 detection

The major challenge during presence/absence testing in molecular diagnosis is determinate the cut offs of testing. In this work, we verify using commercial templates of N gene the LoD and Cut offs of each N1 and N2 primers sets (supplementary figure 1). Both N gene targets were able to detect 5 genome copies (GC) per microliter of reaction with Ct value with N1 (34,28±0,6841) and N2 (34,18±0,5382). In recent work, mock group usage shows high Ct values during US CDC N molecular test, suggesting the importance of establishment of different cut offs that are suggested by standard protocols that recommend 40 Ct value (Liu et al. 2020). Herein, we verified the reproducibility of high Ct values in patient samples using N1 and N2 US CDC primer sets (figure 2A). The use of many targets is justified by frequency of alteration in the viral genomes that can reduce the capacity of primer sets to detect the virus, but at same way is responsible to need more sophisticated apparatus to do diagnosis (Liu et al. 2020). We detect no mutation in N gene in the Latin American genome sequences of SARS-CoV-2 in the regions targets of US CDC primers set (supplementary figure 2). In addition, we identified nonspecific amplification using both primers set (figure 2B-C), which can be explained by annealing primers in different human genome regions (supplementary figure 2). Our data suggest that traditional PCR method and acrylamide gels can alternatively be used in remote locals with poor access to molecular tools.

**Figure 2.**
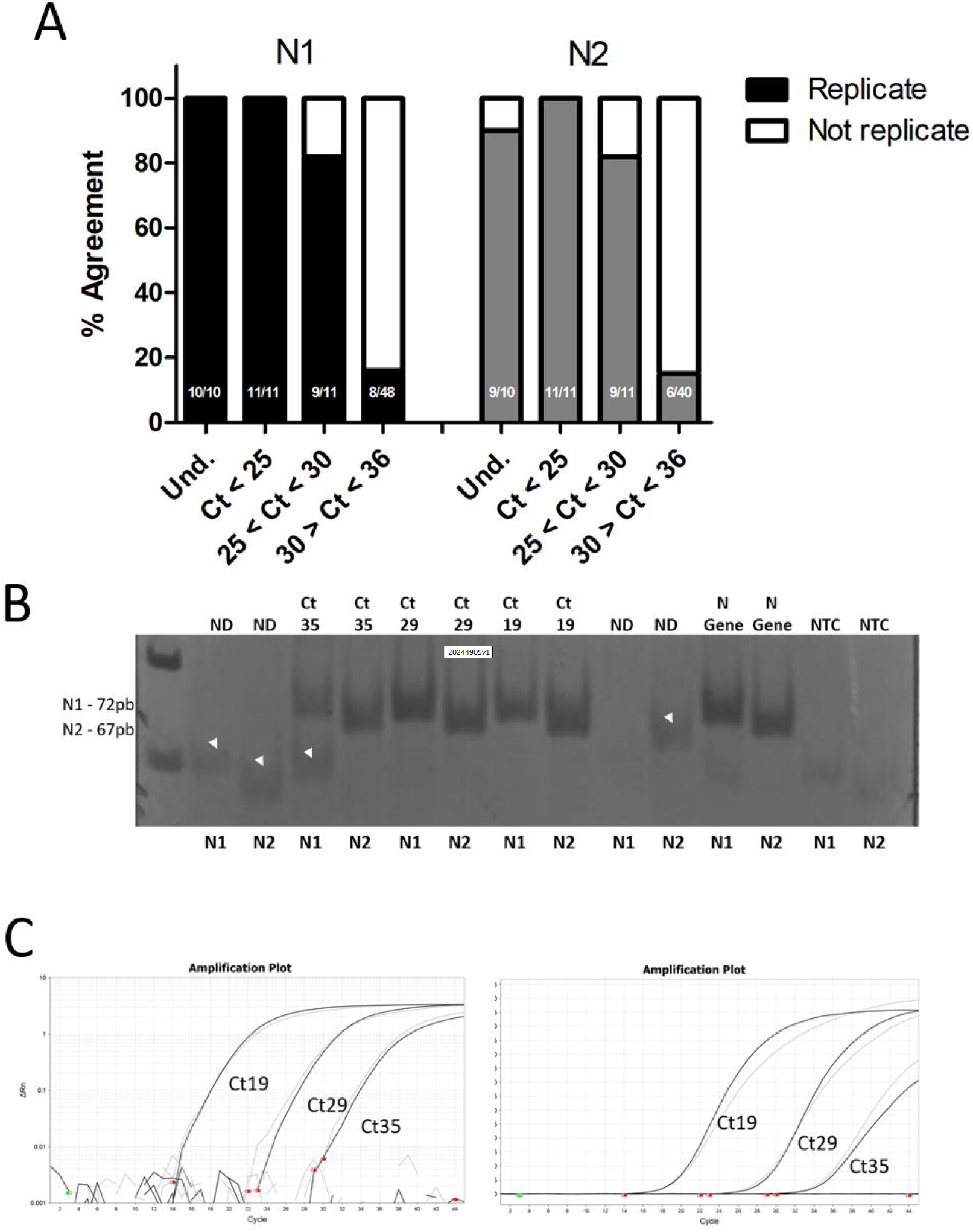
Checking N1/N2 accuracy by Ct value using RT-PCR and polyacrylamide gel. Patient samples were completely reanalyzed to check the accuracy to N1 and N2 gene targets. (A) Contingency graphs display percentages of agreement between two testing in different cycle threshold (Ct) groups. Representative (B) polyacrylamide gel and (C) amplification plots in log (left panel) and linear (right panel) representations. Arrowheads indicate nonspecific amplicons. Und. - undetermined Ct.

### High Ct value has low predictive value to diagnosis

In order to verify the specificity of the US CDC test, we evaluated samples from the same patient that were collected between 2 and 6 days after the first exam and we found that only 32% of the samples were negative, maintained the Ct value and another 33% reduced the Ct value (figure 3A). The ROC curve analysis for patients with Ct above 33 that were doubly positive revealed a low sensitivity (value) and specificity of the test (value) in samples with Ct above 33 (figure 3B). Thus, our data suggest that samples tested with Ct close to the detection limit have a low predictive value and should not be considered for diagnosis before collecting a new sample and performing a second confirmatory test.

**Figure 3.**
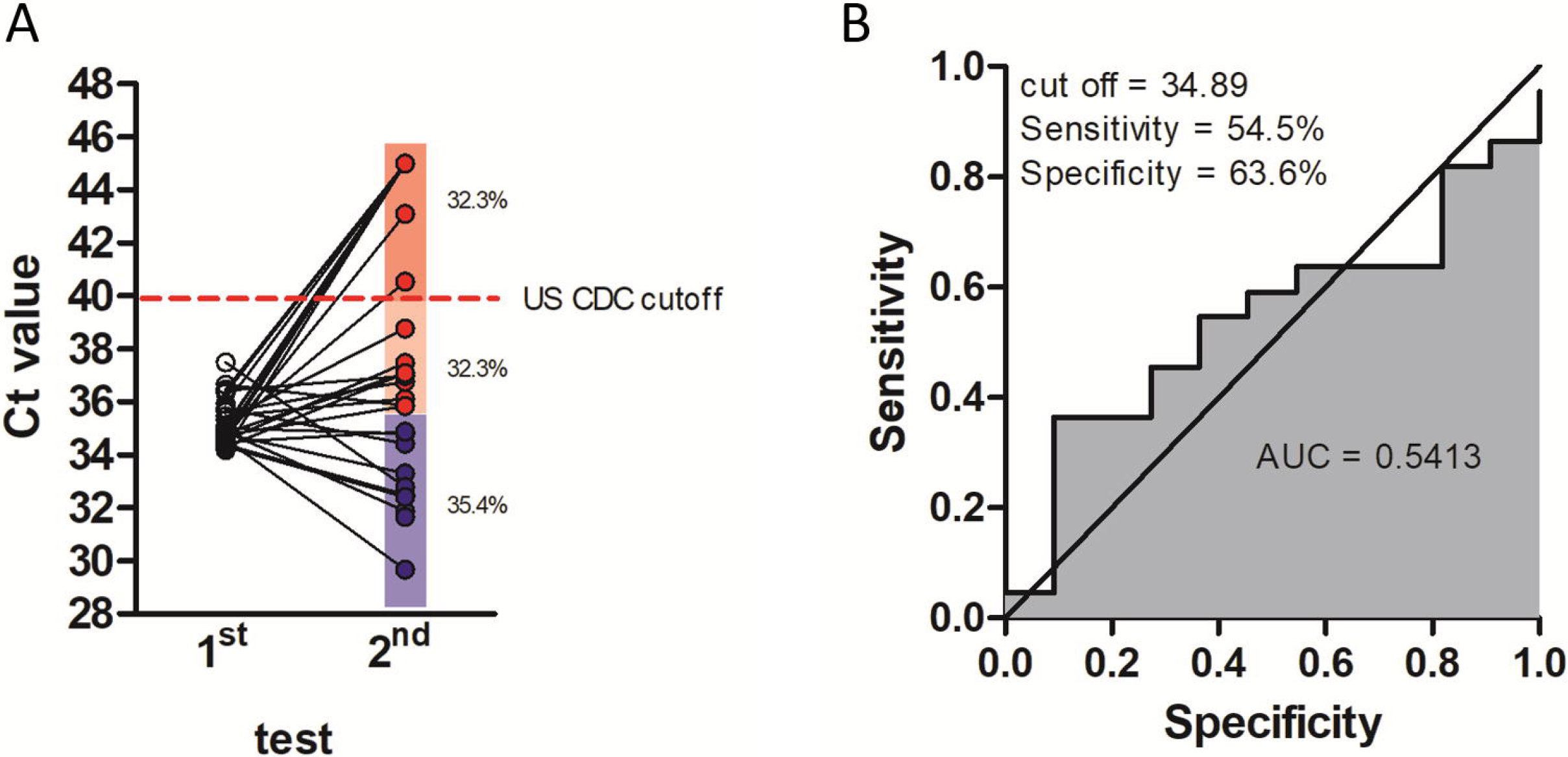
Double testing analysis of high Ct value samples of patients. (A) Patient samples were completely reanalyzed after new patient’s testing until six days after first test. Blue dots indicate reduction and red dots indicate an increase of Ct value after second test. Red dashed line indicates the US CDC cut off to positives samples. (B) A Receiver Operator Characteristics (ROC) curve analysis was performed to assess the sensibility and specificity in the comparison between double positive samples and just first positive sample (presumptive negative). AUC: Area Under Curve.

## 4. Discussion

Analysis of the performance of tests for the detection of the SARS-CoV-2 virus has been carried out worldwide (Liu et al. 2020, Vogels et al. 2020)^’^. The performance of primers and probe sets for virus detection has been evaluated, but there are still few studies that assess the sensitivity and specificity in the critical range of detection of the RT-PCR technique (Mathuria et al. 2020). Among the most sensitive primer and probe sets are those for target N available by the US CDC (Vogels et al. 2020, Etievant et al. 2020)^’^. In this study, we evaluated the performance of the two sets of primers and probes used by the CDC and identified that both presented the same diagnostic performance, suggesting that only one of the targets could be used in the molecular diagnosis of COVID-19, as with other tests that use just one molecular target reducing costs (Pizzol et al. 2020, Mathuria et al. 2020). Liu et al. 2020 evaluated the performance of primer and probes sets from different RT-PCR diagnostic kits for COVID-19, finding results similar to those of our study (Liu et al. 2020). Here, we assessed the performance of the US CDC primer set on sputum and swabs patient samples and noted a similar performance between the two N1 and N2 targets for virus detection. Some studies have compared the sensitivity between specimens for the detection of SARS-CoV-2 and identified that saliva samples may have a similar or superior sensitivity to swab samples (Güçlü et al. 2020, Wyllie et al. 2020).

The RT-qPCR is the gold standard technique for detecting the SARS-CoV-2 virus and it has been used to validate alternative diagnostic methods for COVID-19 (Pizzol et al. 2020, Mathuria et al. 2020, Ai et al. 2020). However, data that assess the sensitivity and specificity of the technique for the higher Ct ranges are still scarce (Mathuria et al. 2020). In this sense, a study evaluated the Ct value of health workers who underwent two tests and found an increase in the Ct value in an interval of 21 days between exams (Cariani et al. 2020). Most commercial diagnostic tests recommend that Ct values below 40 be considered as the cutoff point for a positive diagnosis for coronavirus (Liu et al. 2020, Vogels et al. 2020, on behalf of the SARS-CoV-2 Foch Hospital study group et al. 2020). However, studies that evaluated the detection limit, as well as, our study have shown a low predictive value of RT-qPCR in samples with Ct above 35 using the US CDC protocol (Liu et al. 2020, Vogels et al. 2020). In this study, we found that patients with Ct samples above 33, when retested in a short period of time, may have the test result drastically altered, suggesting that high Ct values have a low positive predictive value. Due to the COVID-19 pandemic and the growing need for molecular testing, many laboratories have not been able to adequately assess the efficiency of the tests made available for use and they are using Ct value below 40 as positive diagnoses for SARS-CoV-2. We identified, by repeating two or more times the sample extraction with high Ct, that these samples have low reproducibility, suggesting the need for patient return to completely repeat the US CDC test for greater diagnostic security. We identified that the low reproducibility of samples in the Ct range above 34 for US CDC primer sets could happen for 3 reasons: (i) cross contamination of the samples during processing; (ii) low viral load in the samples due to the final or initial stage of infection (iii) presence of low viral load close to the detection limit of the technique.

Few cases of reinfection have been reported in different countries around the world (Prado-Vivar et al. 2020, To et al. 2020, Van Elslande et al. 2020, Tillett et al. 2020). The reinfection data report a case of mild infection with low viral load and high Ct value followed by a period without positive serology for SARS-CoV-2 infection and a second infection with high viral load and severe symptoms and followed by serology positive (Prado-Vivar et al. 2020, Tillett et al. 2020). In view of the data in this study, we consider that the authors should exercise caution in stating cases of reinfection based on high Ct values in either of the two episodes reported in the same patient. In addition, a definitive study on humoral response demonstrated a robust long-term production of neutralizing antibodies against SARS-CoV-2 in patients infected only once (Wajnberg et al. 2020). Evidence of reinfection should take into account cases in which there was a clear viral load in both episodes and could rule out the presence of cross-contamination between samples during the analyzes, which could explain both the high Ct values between the samples and the genetic diversity observed. Then, any case report presented viral load below Ct value 30 in both episodes of infection should be considered reinfection (Prado-Vivar et al. 2020, To et al. 2020, Van Elslande et al. 2020, Tillett et al. 2020).

Together, our data show that both N1 and N2 sets of probes and primers can be used individually for the diagnosis of COVID-19. In addition, we found that the RT-qPCR technique involving US CDC primers should be used with caution in the diagnosis of patients whenever the Ct values are close to the detection limit established by each one laboratory services. We recommend that samples with Ct above the detection limit be re-extracted and reanalyzed by an alternative protocol and in cases of doubt, a new examination must be performed on the patient before the diagnosis can be released as positive. The data from this and study has an impact on the interpretation of future data about the COVID-19 pandemic and on the conduct of sample analysis using the N gene as a target.

## Data Availability

The authors confirm that the data supporting the findings of this study are available within the article [and/or] its supplementary material.

## 6. Contributors

Conceived and designed the experiments R.C.K., L.P.R.V., J.B.L., T.A.S. Data collection: L.V.G.K., L.P.R.V., L.G.B., R.C.K., J.B.L., T.A.S; Analyzed the data: R.C.K., L.P.R.V., J.B.L. T.A.S.; Contributed materials/analysis tools: L.V.G.K, M.H.F.K, R.C.K., J.B.L., T.A.S.; Wrote the paper: J.B.L., T.A.S., R.C.K.

## 8. Acknowledgment

We thank the Secretariat of Epidemiological Surveillance of the city of Barreiras and the West Hospital for providing data on patients treated in the municipality. We also thank Rosilene, Iole and Marlan for technician support.

